# Autocatalytic Model for Covid-19 Progression in a Country

**DOI:** 10.1101/2020.04.03.20052985

**Authors:** Anatoly Chernyshev

## Abstract

Herewith we present a computational model for the forecasting of cumulative diagnosed cases of Covid-19 pneumonia within a country. The only explicit parameter of the model is the population density. The implicit parameter is a moving average ambient temperature, currently integrated into the kinetic constants. Other finer details pertaining to the mechanism of the pathogen SARS-CoV-2 spread within a given region are implicitly manifested in the exponent parameters derived from the non-linear fitting of the published data on Covid-19 occurrence. The performance of the model is demonstrated on a few selected countries, and on the Diamond Princess cruising ship outbreak. The model might be used as an aiding tool for the policy makers regarding the decisions on the containment measures and quarantine regime required.

## Introduction

The recent spread of freshly emerged pneumonia viral pathogen SARS-CoV-2 originating in Hubei province (China) has caused a great havoc across all regions of the world. First time in the civilization history quarantine measures of different magnitude were implemented in almost all countries, and passenger traffic ceased. The governmental reactions to the disease spread ranged. South-East Asia countries were tending more to strict martial-law style restrictions. Some of the Northern Europe governments were trying to promote “herd immunity” concept, where the vulnerable members of the society (e.g. the elderly and immuno-compromised) were isolated, while the others were allowed to be with no restrictions. Allegedly, this strategy was to eventually terminate the disease spread due to most of the society would be immune.

The important result of all these diverse actions is that each country happened to be in a sufficient isolation from the others, maintaining its specific climate and social conditions.

It is hardly possible to account for all miniscule factors contributing to the pathogen spread in each case, so that one needs certain simplifications in order to be able to forecast the disease progression.

The following factors are drawing immediate attention:

1. The diagnosed cases appear to be able to reach a plateau, without spreading to all population (as witnessed by the examples of China and Diamond Princess ship).
2. There is an indication that the disease spread is much slower in hotter climates and in certain humidity levels. A clear example is India and most of Africa, where the cases are very low compared to the population numbers. Indeed, recent studies have shown that the temperature optimum for the SARS-CoV viruses stability in the environment is around 4°C, yet the humidity also have significant impact [1, 2].
3. A relatively lengthy incubation period (5–14 days [3]) combined with about tenfold rate of undiagnosed cases [4] compared to the reported ones signals that the virus is struggling reproducing in humans. In other terms, it is fast to spread, but not thermodynamically stable once inside the body.

## Model

These observations fit well into a range of processes, which could be described by the kinetics of an autocatalytic chemical reaction. The corresponding model and its outcomes is presented below.

The data for the model were obtained from a daily updated resource at European Centre for Disease Prevention and Control (https://www.ecdc.europa.eu/en/publications-data/download-todays-data-geographic-distribution-covid-19-cases-worldwide).

There was only one country so far, which has clearly demonstrated the total number of diagnosed Covid-19 case to reach a plateau, China (Fig. 1).

**Fig. 1.**
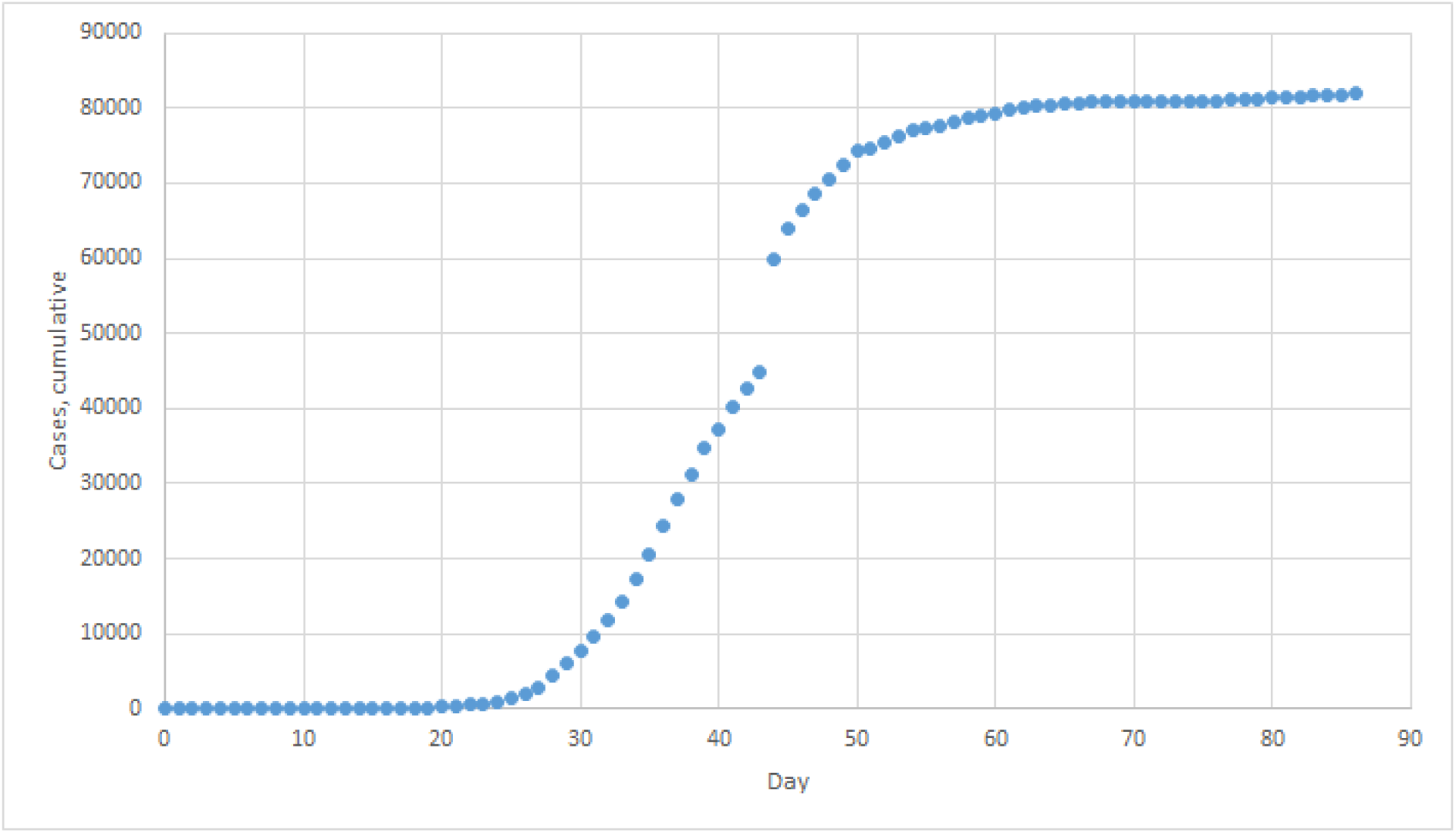
The dynamics of cumulative diagnosed cases in China, from 31/12/2019 to 26/03/2020.

Assuming that would be a general scenario other countries will follow, the starting points of the model were derived from this case.

The S-shaped curve on Fig. 1 is typical for autocatalytic reactions, when the rate of the reaction is accelerated by its product:

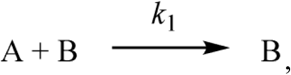

Where *k*_1_ is the rate constant in terms of standard chemical kinetics. Indeed, if we take the number of healthy individuals for A, and the number of infected individuals for B, this reaction seems to be suitable for epidemiological modelling. We need some infected people to infect the healthy ones, and the more of sick people are around the faster it will happen:

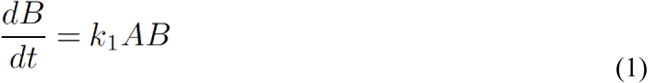

The important note is that from this point on, the values of A and B will be expressed in units of population density, that is the number of (un)healthy individuals divided by the country’s land area (km^2^). The population density shall serve as a proxy for chemical concentration in standard kinetic modelling.

In the simple case above the process will go uninterrupted until no more healthy people available. As shown on Fig. 1, the infection has stopped at approximately 80,000 individuals, which is a tiny fraction of China’s 1.4 billion population. When such phenomena are observed in chemical processes, this is an indication of an existing equilibrium between the forward and reverse reaction, the latter proceeding with a separate rate constant *k*_2_:

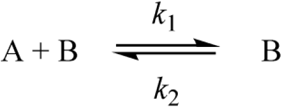

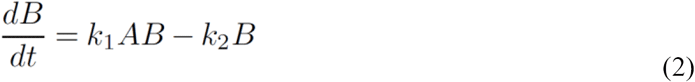

We have attempted first to fit the observed Covid-19 dynamics in China using equation (2), but the results were not satisfactory.

The next step was to test a general case of the equilibrium above by introducing “stoichiometric coefficients”^1^ *a*_1_, *b*_1_, *b*_2_:

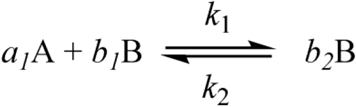

Then the rate is expressed as following:

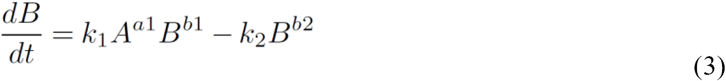

Since the total number of healthy and sick individuals equals to the country population, A + B equals the population density, which is constant for a given country, *d*_*P*_ = total population/land area. Then we obtain the final expression for the infection rate:

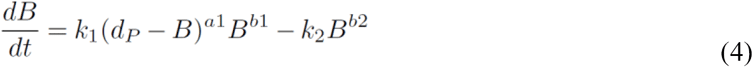

The parameters *k*_1_, *k*_2_ are the “reaction rate constants”, specific for each country. They potentially allow one to account for the effect of temperature on the disease spread using the Arrhenius equation:

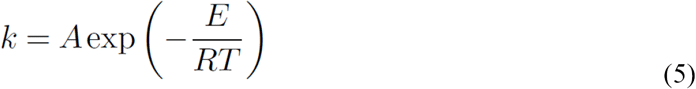

Which for the epidemiology purposes could be simplified to

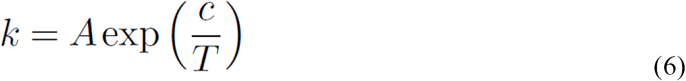

Where A and c are some constants specific for the region.

The tree exponents shall incorporate implicitly the specifics of the mechanisms of transmission pertaining to the country, such as:

- Government (un)action (e.g. introduction of quarantine, or “herd immunity”, or else…)
- Local habits and traditions (e.g. wearing masks, mass gathering);
- Local diet (e.g. turmeric-rich diet in India or fast food preferences);
- Local prevailing genotype.

To obtain the model parameters, a subset of published data on each country was used, beginning from some day when the country infection becomes evident (i.e. when the total numbers are still low, but the new cases are reported every day). The experimental rate (day^-1^) *d*B/*d*t is equal then to the number of reported new cases divided by the country area. The simulated rate will be the one calculated from eq. (4) using a test set of the five model parameters (*a*_1_, *b*_1_, *b*_2_, *k*_1_, *k*_2_). The final set of parameters was obtained by minimizing the sum of mean square deviations: 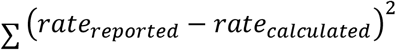 for the specified data range.

## Results

We have finished simulations on ten countries and Diamond Princess liner. Regrettably only China, the liner, and probably South Korea could be used to demonstrate the performance of the model. Other countries were modelled blindly, so that these instances could be proven right or wrong only as the disease propagates. Nevertheless, below we are presenting current results, with some critical remarks.

### 1. China

This is the only country which is allegedly about to complete the infection cycle, with 81620 total diagnosed cases and 1727 remaining active cases (as of April 03, 2020). Therefore it has been the main testing ground for the model. Below we present four modelling outcomes, which differ only in the selection of the data set (red dots on the graphs). Brown dots are the cumulative reported cases, which were not included in the model; blue dashed line is the model’s forecast based on the start day.

As expected, a kinetic system based on a non-linear equation (eq. 4) is quite sensitive to the data selection, and currently there is no universal criteria of how this selection should be done. Some insights on that are given at the end of the manuscript.

However, the main uncertainty of the model is in the plateau height; the bend dates appear to be congruent. Currently “the bend” is defined simply as the date when 85% of the calculated plateau cases reached.

Surprisingly, when only half of the data is used (Fig. 3) the fit was much better on one occasion. This would represent a real-life application of the model, and demonstrates its robustness on the incomplete data sets.

**Fig. 2.**
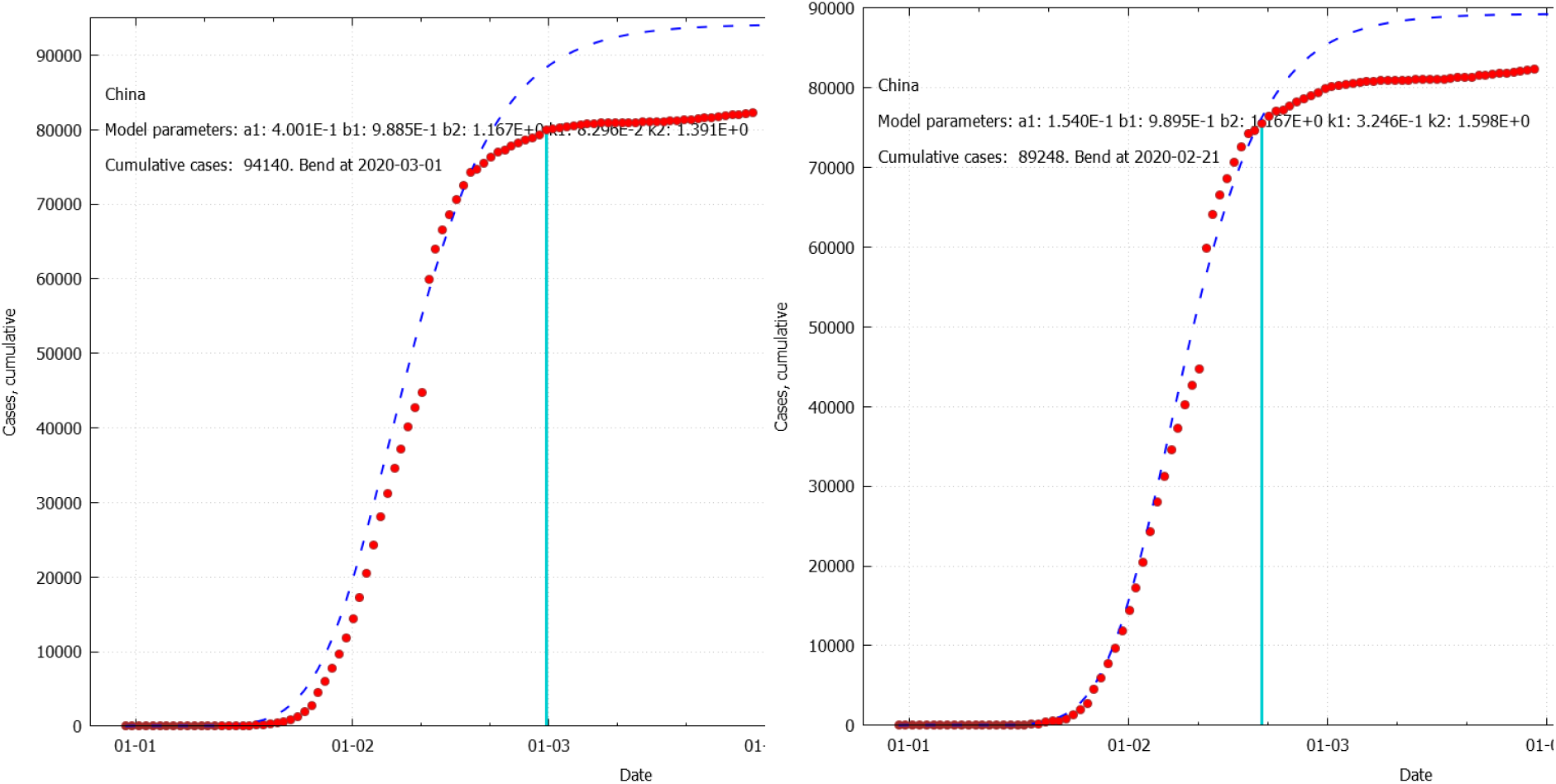
The model for China based on complete data set starting from day 15 (left) or day 21 (right).

**Fig. 3.**
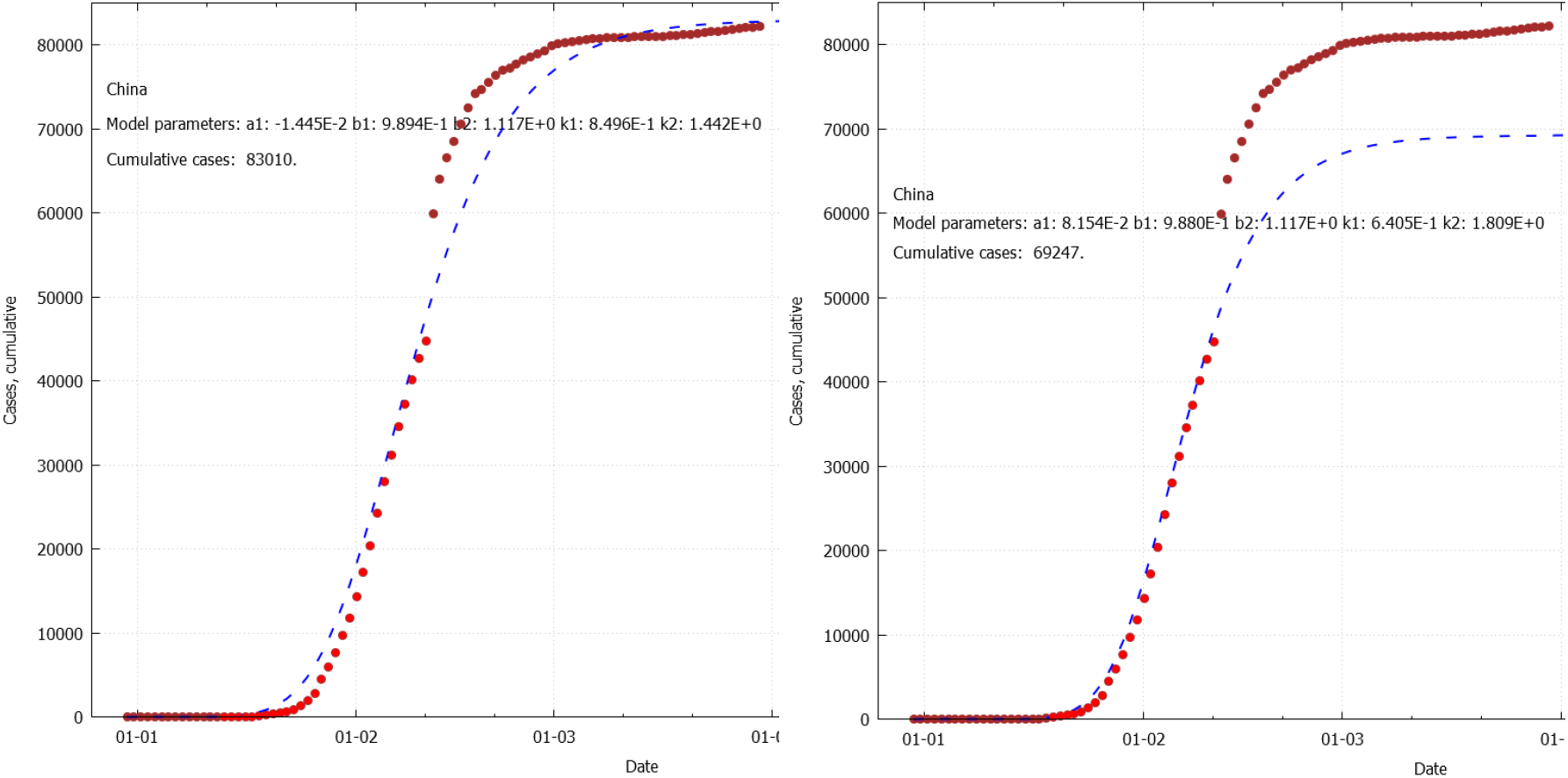
The model for China based on the data set before the big gap in the middle (day 15-44, left; day 21–44, right).

## 2. Diamond Princess cruise ship

Diamond Princess is a cruise ship, which was bound to quarantine in the port of Yokohama, Japan on 5 February 2020 after a passenger has been diagnosed with Covid-19. The epidemiology of the virus outbreak there was well studied in recent publications [5, 6].

Diamond Princess had carried total of 3711 people, which translates into a very high local population density of ∼26500 persons/km^2^. This is roughly 3 times higher than the most populous country, Singapore, and, apparently this has resulted in a steep infection rate. Nevertheless, by the time disembarkation on March 1^st^ 2020, only approximately 20% of people onboard were tested positive for SARS-CoV-2.

Despite a very unusual environment, the model handled this case perfectly well. Most datasets were producing a forecast exactly reproducing the reported dynamics (Fig. 4, top).

**Fig. 4.**
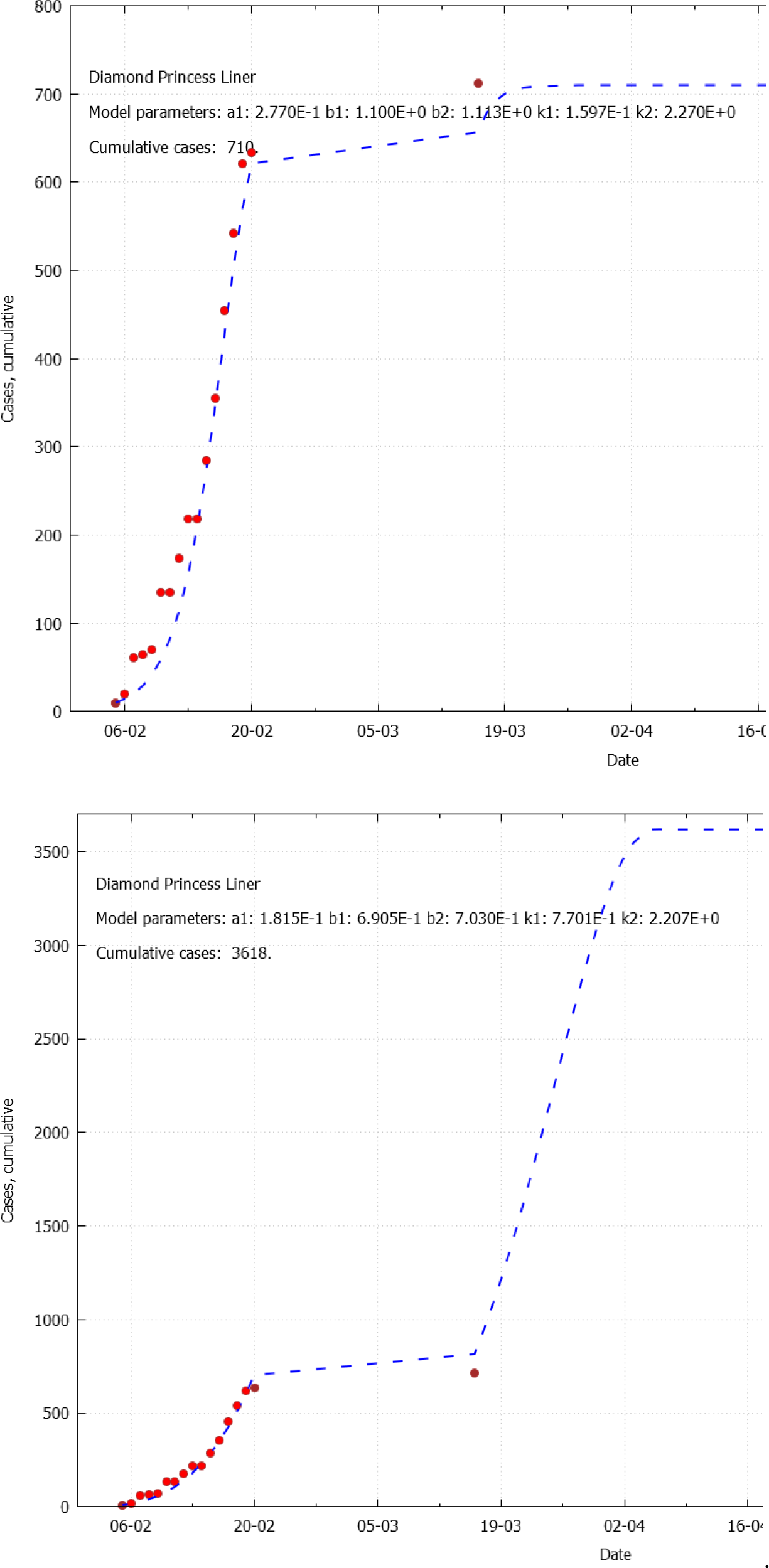
The model for Diamond Princess liner based on day 2-16, top; day 2–15, bottom.

In one particular curious case, if just one extra day was omitted from the model, it predicted a sharp rise in the infection rates, up to 97% of the total passengers (Fig. 4, bottom). We prefer to think of this scenario as an artefact of a non-linear system subjected to unnatural conditions (high population density), but it also highlights the theoretically possible outcomes.

### 3. South Korea

South Korea is another interesting case, with a kink in the S-curve. The model was not able to reproduce this kink, but it was consistent in predicting ∼10000 of total cases, flattening at the end of March 2020 (Fig. 5).

**Fig. 5.**
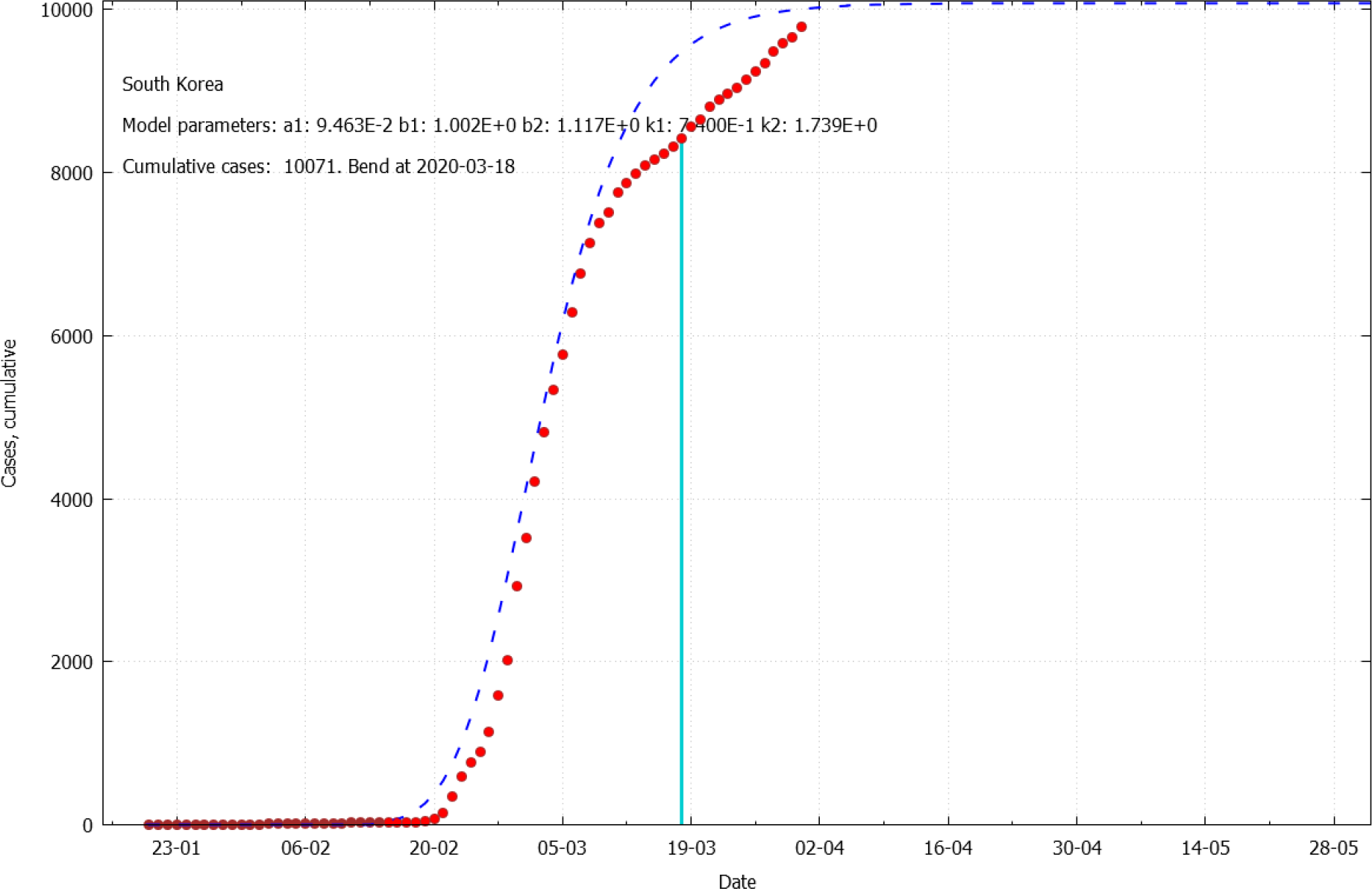
The model for South Korea days 47-92.

It appears that the models might behave quite unpredictably depending on the selection of the start day in the flat initial region, when the fluctuation in the rates is high. Therefore, one has to run the simulation for a range of start dates (say, from the very beginning to the visible bend in the S-curve), and then derive the conclusion based on the most frequent scenarios. As it is demonstrated below, some countries are more stable than other.

### 4. Congruent countries: New Zealand, Italy, Spain

Apparently, for certain countries, which have advanced well up on their S-curve, the model produces stable solutions. The solutions have certain spread in maximal cases and in the bend date, but this spread could be considered acceptable.

New Zealand: the results are very reproducible, with the maximal cumulative cases ranging from 962 to 1271, and mean date for flattening of 05/04/2020 (Fig. 6).

**Fig. 6.**
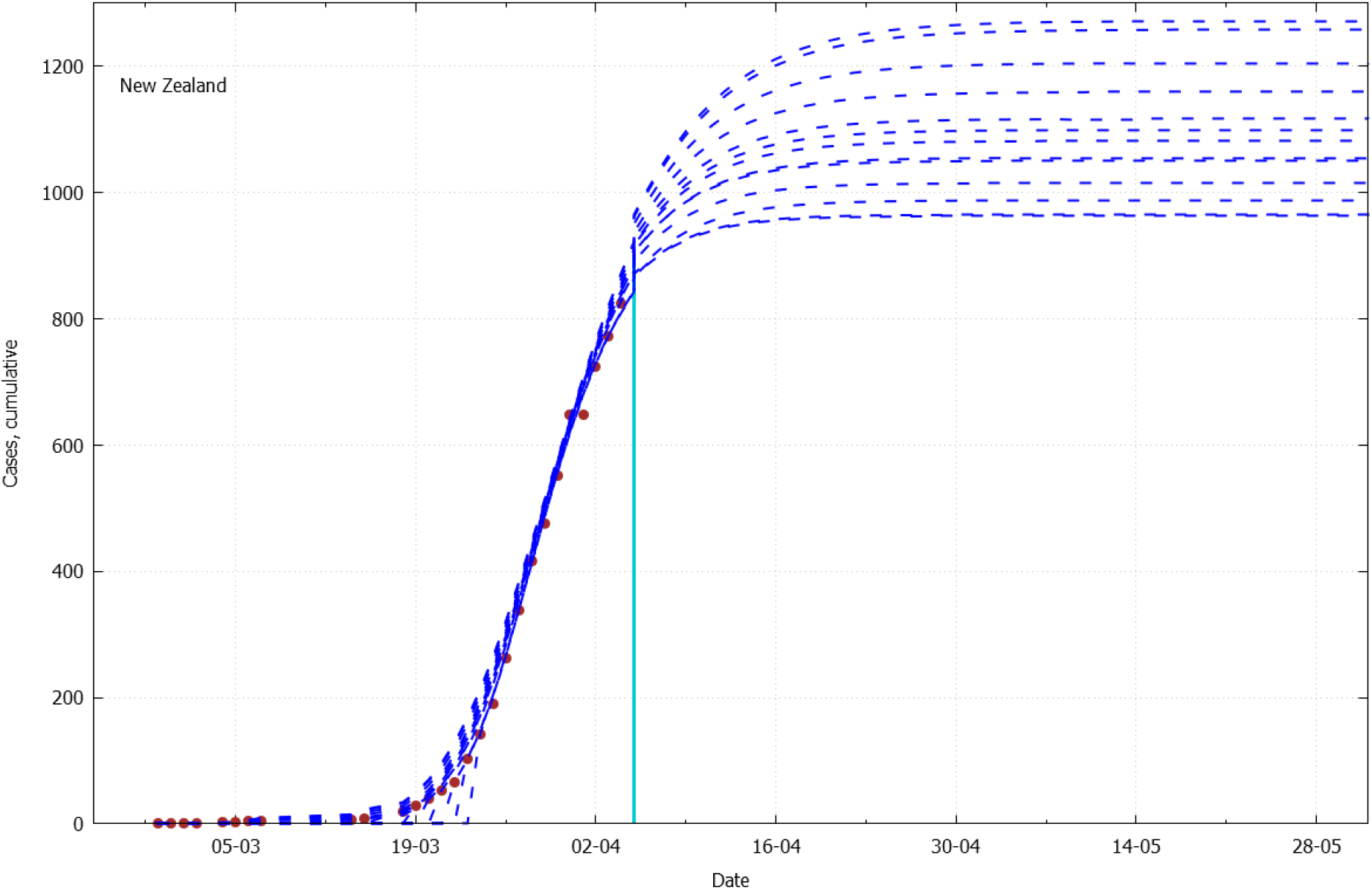
The Covid-19 progression model for New Zealand.

The same for Italy, with maximal cases predicted from 158,000 to 169,000, and with mean flattening date of 08/04/2020 (Fig. 7).

**Fig. 7.**
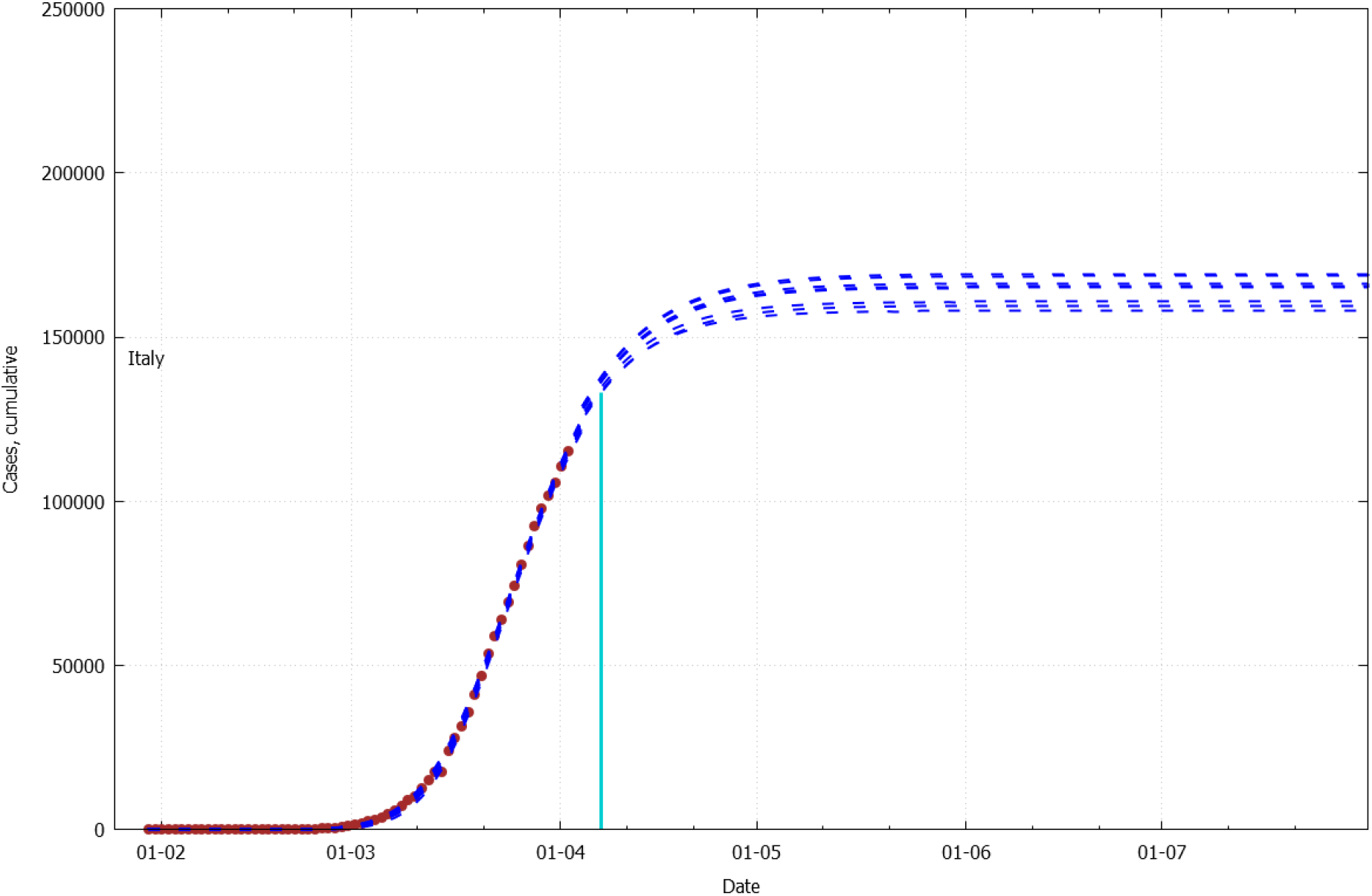
The Covid-19 progression model for Italy.

Spain: 197,000–231,000 total cases; flattening around 14/04/2020 (Fig. 8). Italy and Spain have the same vertical scale for easy comparison, since these two countries had a similar dynamics of Covid-19 spread.

**Fig. 8.**
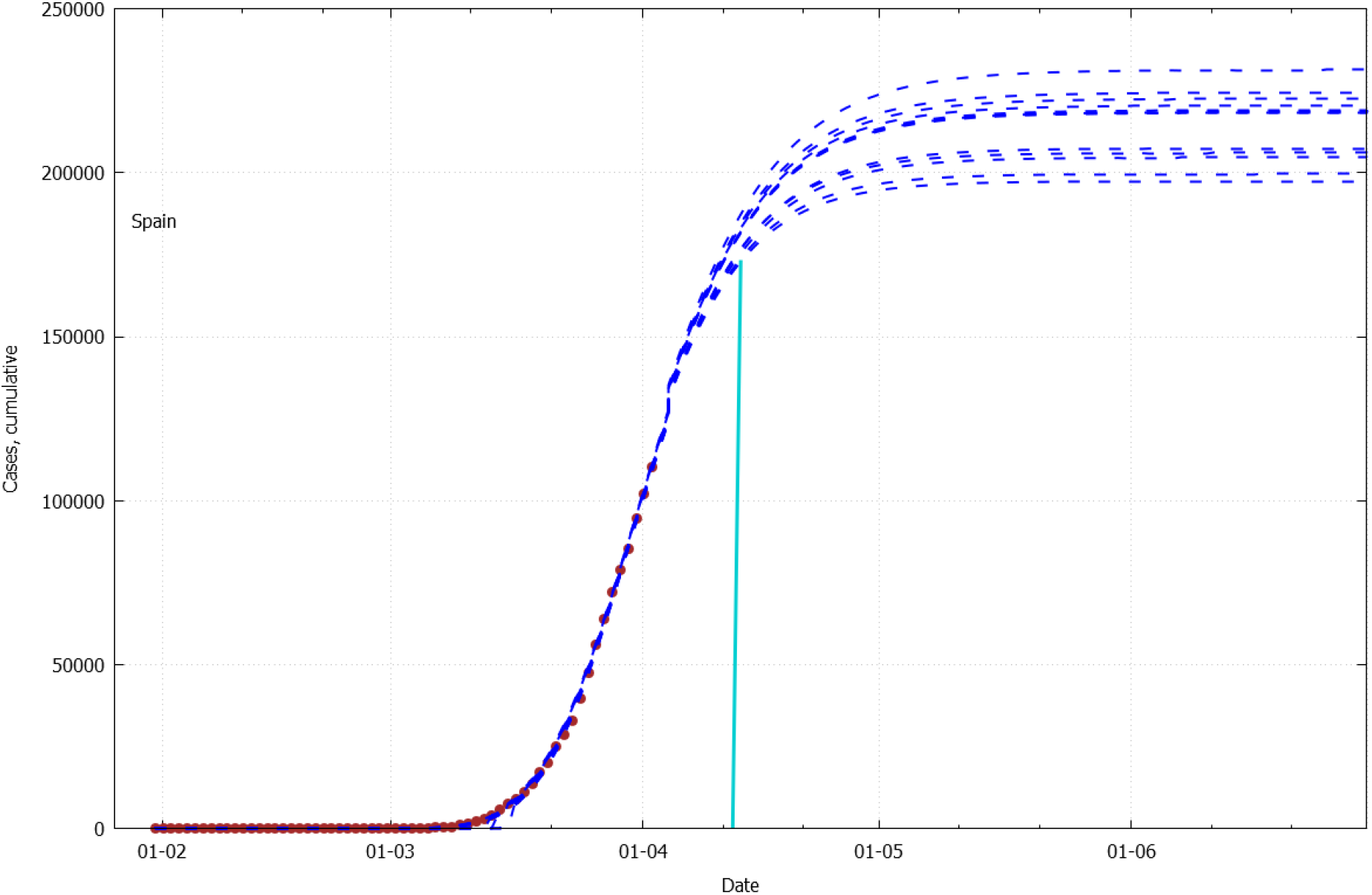
The Covid-19 progression model for Spain.

### 5. Chaotic countries: Singapore, Sweden, India, USA

For the countries, which appear to be in the very beginning (flat portion) of their S-curves, the model demonstrates a very unstable behaviour. For example, for India it predicts the total number of cases anywhere from 13,000 to almost the whole population (Fig. 9). The countries in this category are very diverse in terms of climate, governmental practices, and societal norms. So, for the time being the possible solution would be to wait until the number of cases in each such country yields a convergent forecast. The good news is that for each of the chaotic scenarios, the bending point is in not-so-distant future (for India it ranges from the beginning of June to middle of October 2020).

**Fig. 9.**
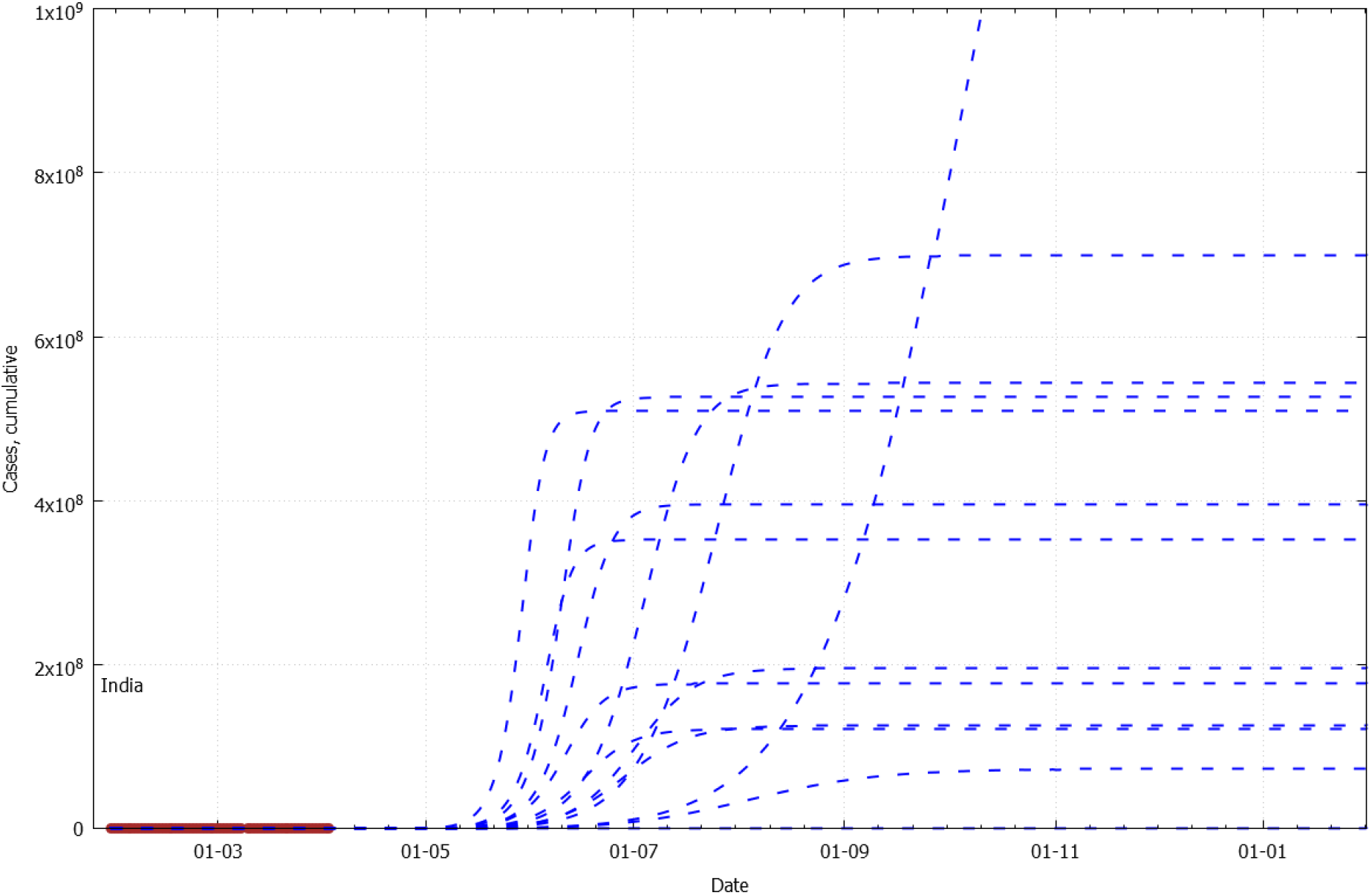
The Covid-19 progression model for India.

Fig. 10–12 show similar pattern for Singapore, Sweden, and the USA.

**Fig. 10.**
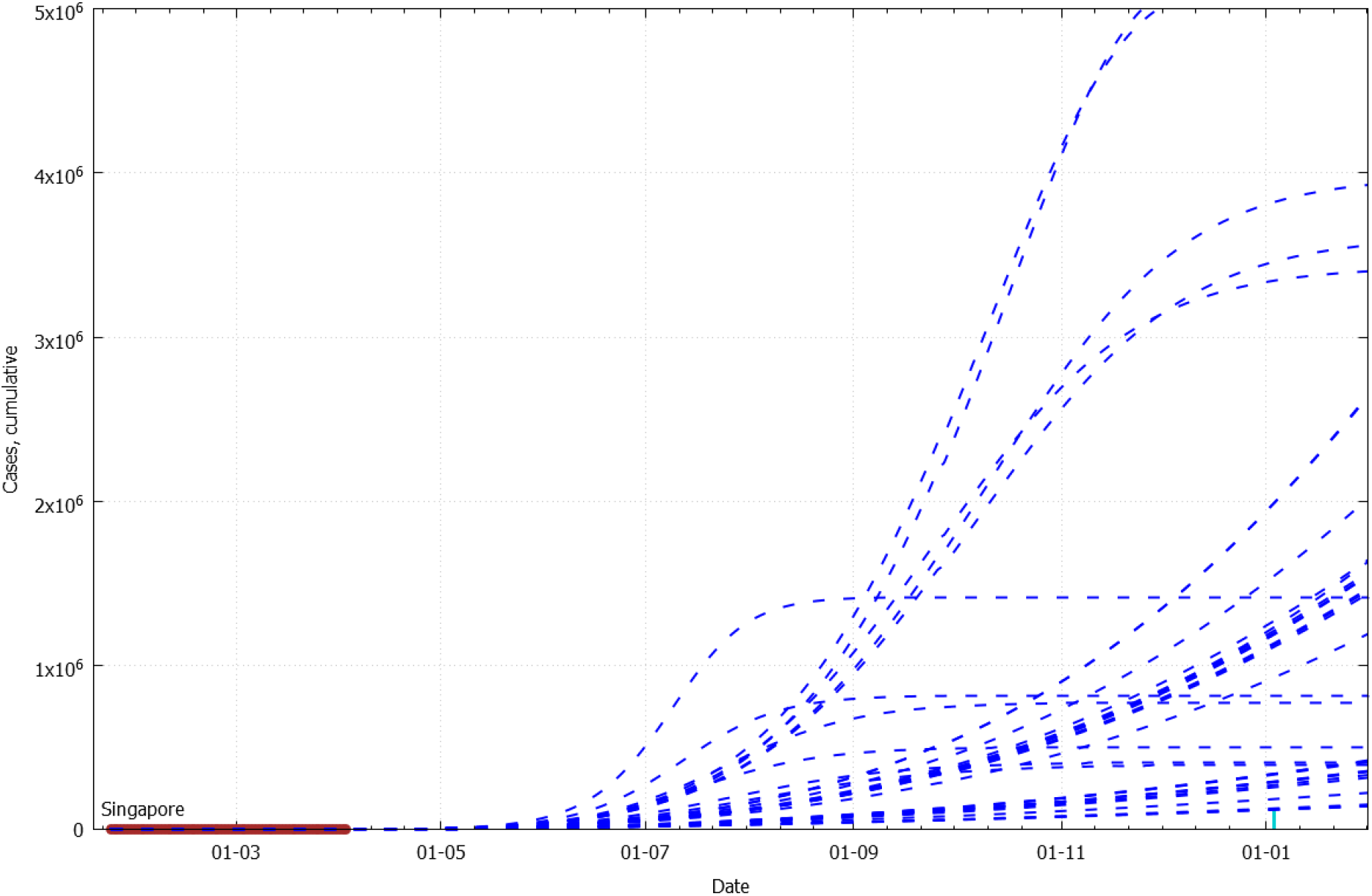
The Covid-19 progression model for Singapore.

The possibility to detect the transformation of a country’s regime from chaotic to stable is demonstrated below (Fig. 13) using the example of Italy. In the very beginning, the forecasts were completely random until enough data was accumulated. At the end (95 days of data), the model is quite consistent (Fig. 7).

**Fig. 11.**
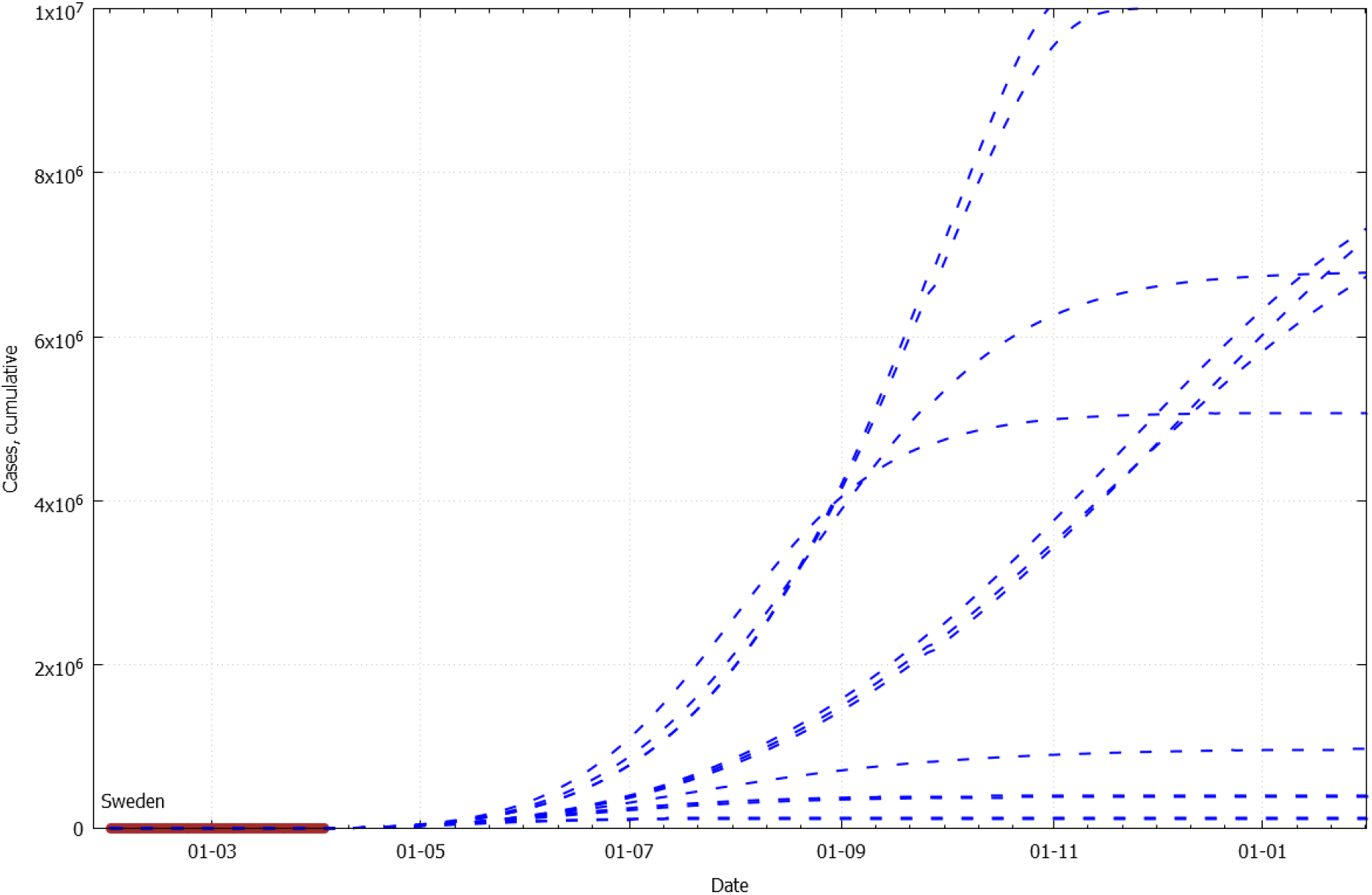
The Covid-19 progression model for Sweden.

**Fig. 12.**
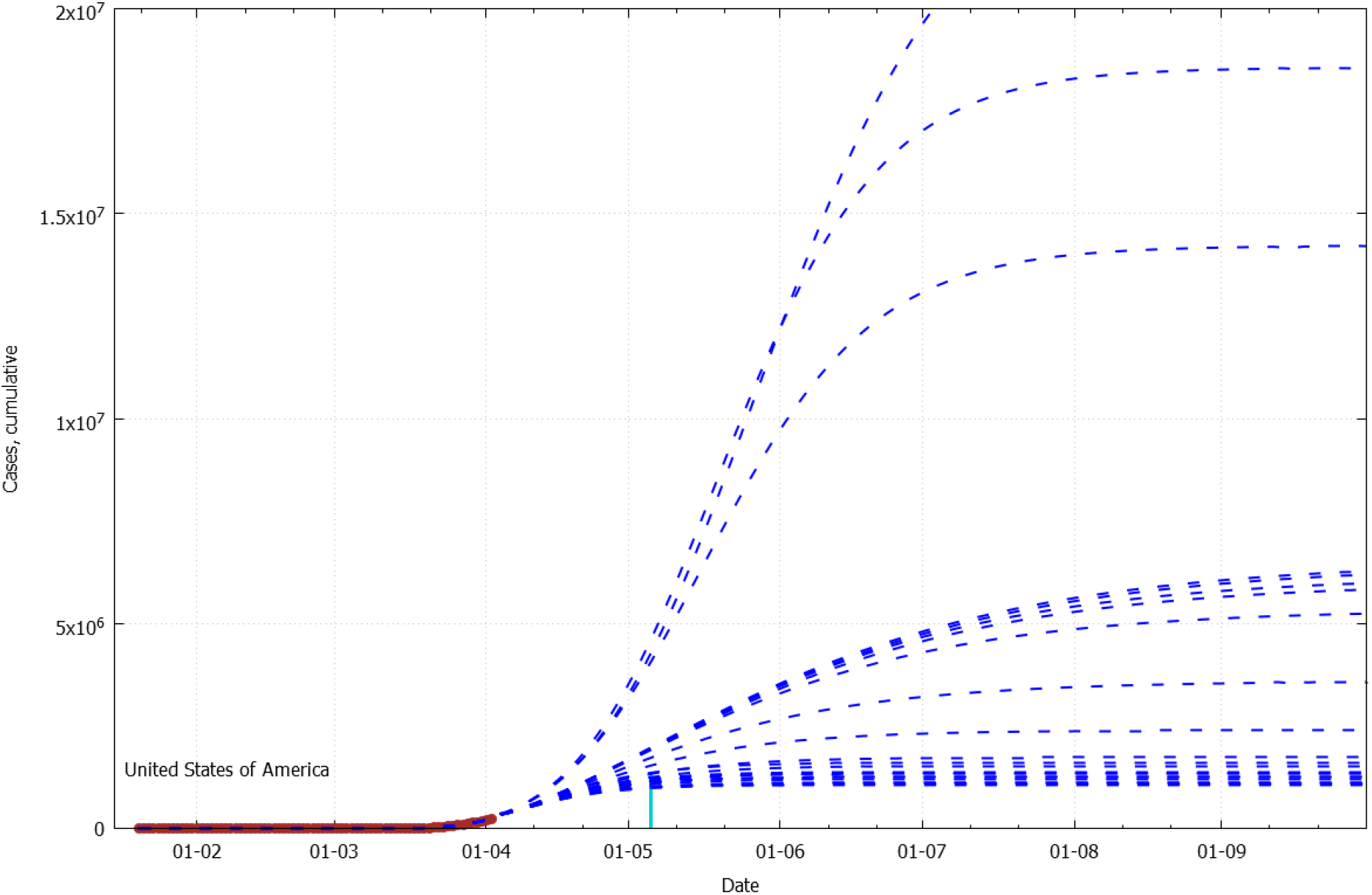
The Covid-19 progression model for the USA.

**Fig. 13.**
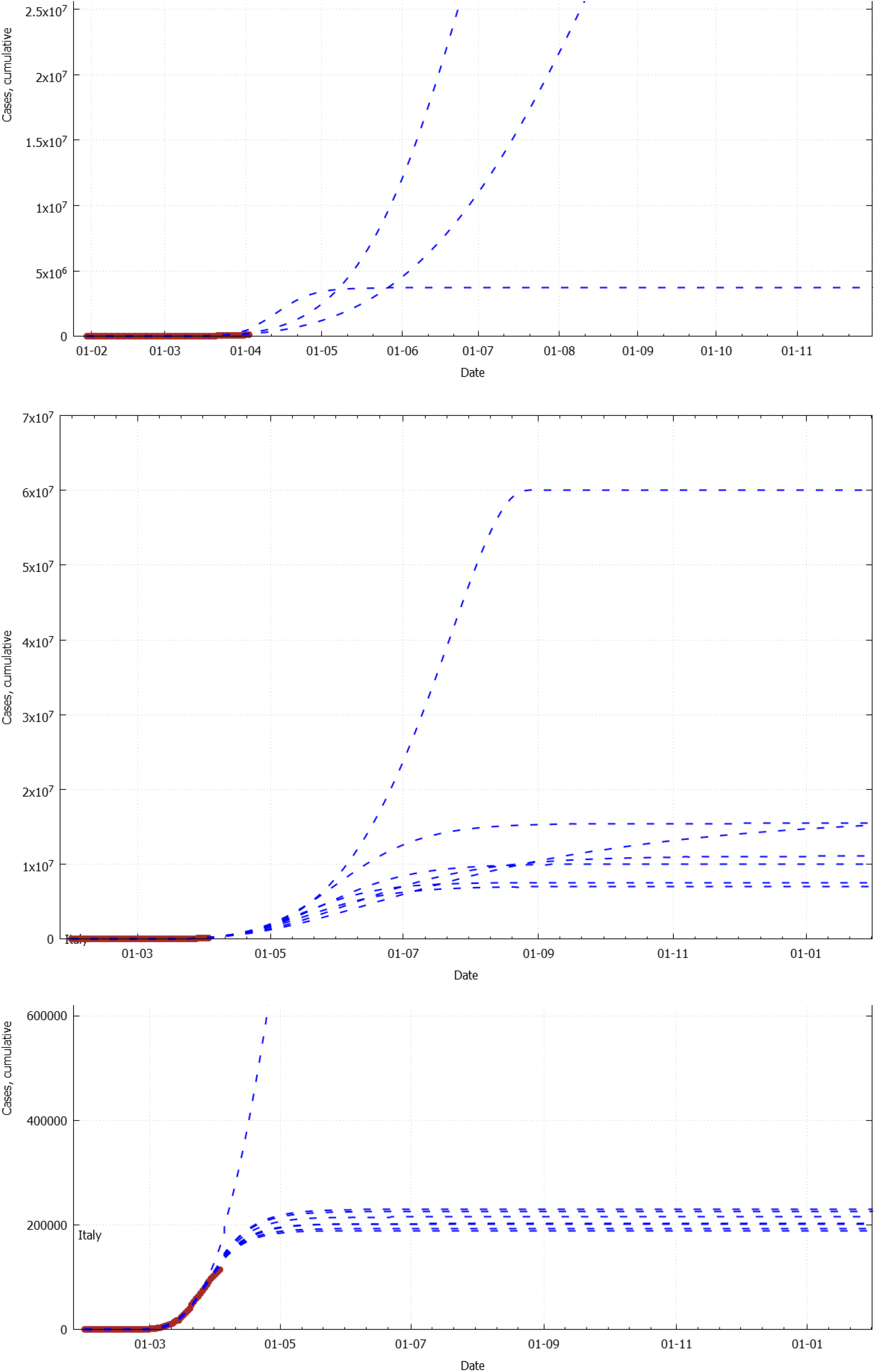
The three stages of model’s chaotic behaviour on the data for Italy. Top: data used up to day 69 (many calculations have failed); Middle: data up to day 75; Bottom: up to day 85 (just one calculation is non-consistent).

This behaviour of the model makes it unusable for the forecast of the infection progression in the countries with low reported cases (notably, most of African countries)

## Conclusions

The presented model for Covid-19 progression has demonstrated good performance on the finished cases (China and Diamond Princess liner). It appears that it is capable to detect the countries, which are well advanced on the S-curve, and close to the plateau. For these countries the model produces stable solutions regardless of the starting conditions. If the solutions demonstrate chaotic behaviour, it might be an indicator that the country is in the very beginning of its local epidemy. General recommendation in this case would be to rebuild the model periodically, as the new cases are reported, until the forecast curves are converged with an acceptable spread.

## Data Availability

The supplementary material contains the results of all calculations presented here, including high resolution graphs, and raw csv data files with the model parameters.
The software, technical details, updates for the model, and forecasts for other countries will be deposited at www.xph.co.nz/index.php/covid-19-progression-model

https://www.ecdc.europa.eu/en/publications-data/download-todays-data-geographic-distribution-covid-19-cases-worldwide

https://www.xph.co.nz/index.php/covid-19-progression-model

## Supplementary

The supplementary material contains the results of all calculations presented here, including high resolution graphs, and raw csv data files with the model parameters.

The software, technical details, updates for the model, and forecasts for other countries will be deposited at www.xph.co.nz/index.php/covid-19-progression-model

## Footnotes

1 We are using quotation marks since the modelling is done by analogy with chemical equilibrium; in the context of pathogen transmission these coefficients shall have a different meaning.

